# Population-level changes in the mental health of UK workers during the COVID-19 pandemic: A longitudinal study using Understanding Society

**DOI:** 10.1101/2021.11.25.21266866

**Authors:** Theocharis Kromydas, Michael Green, Peter Craig, Srinivasa Vittal Katikireddi, Alastair H Leyland, Claire L Niedzwiedz, Anna Pearce, Rachel M Thomson, Evangelia Demou

## Abstract

**Objectives:** The COVID-19 pandemic has substantially affected workers’ mental health. We investigated changes in UK workers’ mental health by industry, social class, and occupation and differential effects by UK country of residence, gender and age.

**Methods:** We used representative Understanding Society data from 6,474 adults (41,207 observations) in paid employment who participated in pre-pandemic (2017-2020) and at least one COVID-19 survey. The outcome was psychological distress (General Health Questionnaire-12; score>=4). Exposures were industry, social class and occupation and are examined separately. Mixed–effects logistic regression was used to estimate relative (OR) and absolute (%) increases in distress before and during pandemic. Differential effects were investigated for UK countries of residence (Non-England/England), gender (Male/female), and age (Younger/Older) using 3-way interaction effects.

**Results:** Psychological distress increased in relative terms most for ‘professional, scientific and technical’ (OR:3.15, 95% CI 2.17–4.59) industry in the pandemic versus pre-pandemic period. Absolute risk increased most in ‘hospitality’ (+11.4%). For social class, ‘small employers/self-employed’ were most affected in relative and absolute terms (OR:3.24, 95% CI 2.28–4.63; +10.3%). Across occupations ‘Sales and customer service’ (OR:3.01, 95% CI 1.61–5.62; +10.7%) had the greatest increase. Analysis with 3-way interactions showed considerable gender differences, while for UK country of residence and age results are mixed.

**Conclusions:** Psychological distress increases during the COVID-19 pandemic were concentrated among ‘professional and technical’ and ‘hospitality’ industries, ‘small employers/self-employed’ and ‘sales and customers service’ workers. Female workers often exhibited greater differences in risk by industry and occupation. Policies supporting these industries and groups are needed.

What is already known about this subject?
Employment has been disrupted by the COVID-19 pandemic and non-pharmaceutical interventions (e.g. national and regional ‘lockdowns’) introduced to control the pandemic. The pandemic has impacted on different occupational groups in different ways and has been linked to substantial deteriorations in mental health.

What are the new findings?
The effect of the COVID-19 pandemic on mental health has been particularly pronounced for those working in professional and technical industries, hospitality, customer service occupations, small employers and the self-employed as well as female workers.

How might this impact on policy or clinical practice in the foreseeable future?
Policies should prioritise support to certain industries, occupations, the self-employed/small business owners, and particular demographic groups (e.g., women in sales and customer service occupations, younger ‘construction’ or non-England workers in ‘Public Administration and Defence’) with high risk.

## Introduction

The COVID-19 pandemic has been linked to substantial deteriorations in mental health (1-3). Work is a well-established determinant of mental health in working-age people, but it has been subject to major disruption. While having a stable and secure job is linked to good mental health, precarious work, income loss and job insecurity have been linked to poorer mental health (4 5). Thus far, occupational health research during the pandemic has largely and understandably focused on the needs of healthcare workers (1 3 6-9). However, the pandemic also poses a major threat to the mental health of the broader workforce.

In the UK and internationally employment has been disrupted by high levels of COVID-19 disease and non-pharmaceutical interventions (such as national and regional ‘lockdown’ measures) introduced to control the pandemic. This disruption has impacted on occupational groups in different ways (1 10-12). For some people, home working allowed employment to continue but was affected by competing domestic responsibilities (e.g., childcare) – often disproportionately affecting women (2 13 14). Others have been unable to work but have retained labour market attachment through the UK Government’s ‘Furlough’ scheme which paid employees at a rate of 80% of their usual salary. In contrast, more precarious workers (e.g., those on ‘zero hours’ contracts) were not well protected by the furlough scheme and therefore at risk of job and income loss.

The pandemic has therefore been a major shock for employment and work, but its health implications remain under-investigated. Links between employment status, job security, work-related stress and mental health are well-established (15-18) and evidence shows the impact of the pandemic on mental health (2 5 19 20). Importantly, impacts are likely to be differential across industries, social class and occupations, as well as across geographical locations such as regions and/or countries, due to differences in work-related factors, such as job demands, control, job insecurity and the protective measures implemented at regional or national levels (i.e., lockdowns, furlough scheme). There is also growing evidence that the economic consequences of the COVID-19 pandemic are particularly negative for young people (21) and are a significant concern for older workers (10). We therefore investigated how the mental health of UK workers changed during the COVID-19 pandemic by industrial sector, social class and occupation, as well as whether any observed changes differed by age, gender, and UK country of residence.

## Methods

### Study population

This study uses the UK Household Longitudinal Study (also referred to as ‘Understanding Society’ and hereafter abbreviated as ‘Usoc’). Usoc is a nationally representative longitudinal household panel study, described in detail elsewhere (22). All adults aged 16+ years in sampled households are invited to participate. Surveys are normally completed over 24 months, with participants re-interviewed annually. Since the COVID-19 pandemic began additional surveys were introduced (23). For this study we use two pre-pandemic surveys (Survey waves 9 and 10/11; data collected 2017-March 2020) and COVID-19 surveys in April, May, June, July, September, and November 2020 (www.understandingsociety.ac.uk). We analysed data from all adults aged 18+ years in wave 9 who were in paid employment (employed, self-employed, both employed and self-employed or in other employment). Participants were included in the analysis if they had complete wave 9 data (2017-2019), participated or not in Wave 10/11 (2018-2020) and participated in at least one of the COVID-19 surveys.

Proxy respondents, those aged less than 18 years old, not in paid employment and those with zero or missing sampling weights were excluded. We also excluded participants with missing data on key variables of outcome, exposures and covariates used in regression analysis (Figure S1). To account for attrition and item missingness, we reweighted our sample to make it representative of Wave 9 participants (Supplementary Material: weighting strategy).

### Outcome

Our outcome of interest was probable psychological distress, defined by ‘caseness’ (score ≥4) in the General Health Questionnaire-12 (GHQ-12) as assessed across eight surveys (two pre-pandemic and six COVID-19 surveys) (24 25). GHQ-12 is a validated screening tool for psychological distress comprising 12 questions (26). Sensitivity analysis was conducted using a cut-off of three or more for GHQ-12 caseness.

### Exposures

Employment-related exposures of interest were industry, social class, and occupation. Industry was classified using the UK Standard Industrial Classification (SIC) sections (27). Overall, 16 industrial sections were included after adapting the 21 original sections (Table S1A) (27). Industry for COVID-19 survey participants was assigned based on the June and July surveys and imputed backwards and forwards to all valid baseline participants. Where observations were still missing, industrial sector was imputed from available wave 10/11 data, and finally if still missing wave 9 data were used. We tested the assumption that industry remains relatively stable across surveys and over a period spanning a one and two wave difference (pre-pandemic) showed that overall reported industry was stable, with 92.4% on average remaining in the same sector (Table S2).

The National Statistics Socio-Economic Classification (NS-SEC) was used for social class categorisation (28). Social class data were carried forward from pre-pandemic to pandemic surveys, assuming that social class is relatively time-invariant. Transition analysis showed that on average 88.3% of participants remain in the same social class between pre-pandemic surveys.(Table S2).

NS-SEC is a theoretically-informed, composite measure that partially captures employment relations and conditions (e.g. wage vs salary, prospects for promotion, and levels of autonomy) (29). Therefore, we included occupation based on the Standard Occupational Classification (SOC) 2010 nine major groups as another exposure to fully capture occupational effects. Occupation was not available in any of the COVID-19 surveys, so we carried data forward from the pre-pandemic period, assuming stability. Overall, occupation remained relatively stable with 87% staying in the same occupational group during pre-pandemic surveys (Table S2).

### Covariates

We adjusted for potential confounders that might differentially affect change in mental health across employment groups. These were age as a continuous variable along with its quadratic term, gender (male/female), UK country of residence (England/non-England (i.e., Northern Ireland, Scotland, or Wales)), race (white/non-white) and employment type (employed/all other employed (i.e., self-employed, both employed and self-employed or in any other type of employment)).

### Statistical Analysis

Sample characteristics were summarised using frequencies and proportions for the pre- and pandemic periods. We then fitted three mixed–effects generalised logistic regression models with a random intercept for each individual participant, to assess the odds of GHQ-12 caseness by exposure groupings (industry, social class, and occupation). All estimations are expressed as odds ratios (ORs) and represent the population average for each exposure variable. Predictive margins and average marginal effects in terms of probabilities were estimated to provide an indication of the absolute change in prevalence of GHQ-12 caseness in the pandemic versus pre-pandemic period for each exposure. In all models, the pandemic impact is represented by the interaction between a binary variable for the pre- (surveys 9 and 10/11 combined) and pandemic period (April to November surveys) and a categorical variable for industry (Model 1), social class (Model 2) and occupation (Model 3). Constituent terms of each interaction are also included in the model. Therefore, the interaction between the variable that distinguishes pre- and pandemic periods, and our three exposure groupings shows how much greater (OR>1 in multiplicative terms or ΔP(Y=1)>0 in additive terms) or lower (OR<1 or ΔP(Y=1)<0) the excess risk for GHQ-12 caseness associated with the pandemic was for each exposure category separately.

We conducted further analysis by estimating 3-way interactions between each exposure variable, the binary variable that represents pre and pandemic periods and (a) UK country of residence (England/non-England); (b) gender (male/female); and (c) age group (younger-under 50/older worker-over 50), respectively. These models included the same outcome, exposure, and predictor variables and were adjusted for confounding as in Models 1-3 above.

All analysis were conducted in Stata 17.0. Understanding Society has ethical approval granted by the University of Essex Ethics Committee and further approvals were not necessary for this secondary data analysis.

## Results

Our main analytical sample (for Models 1 and 2) comprised 41,207 observations (pre-pandemic period=12,192, pandemic period=29,015) across 6,474 individuals. Analysis by occupation (Model 3), included 27,110 observations across 4,485 individuals (Table 1, Figure S1, Tables S4-S6), due to missingness in the occupation variable. Across the final sample, 57.9% were female and mean age was 48.17±11.33 years. Overall, 8.05% were non-white and for most (81.2%) country of residence was England. Most participants (84.5%) were employees.

**Table 1:**
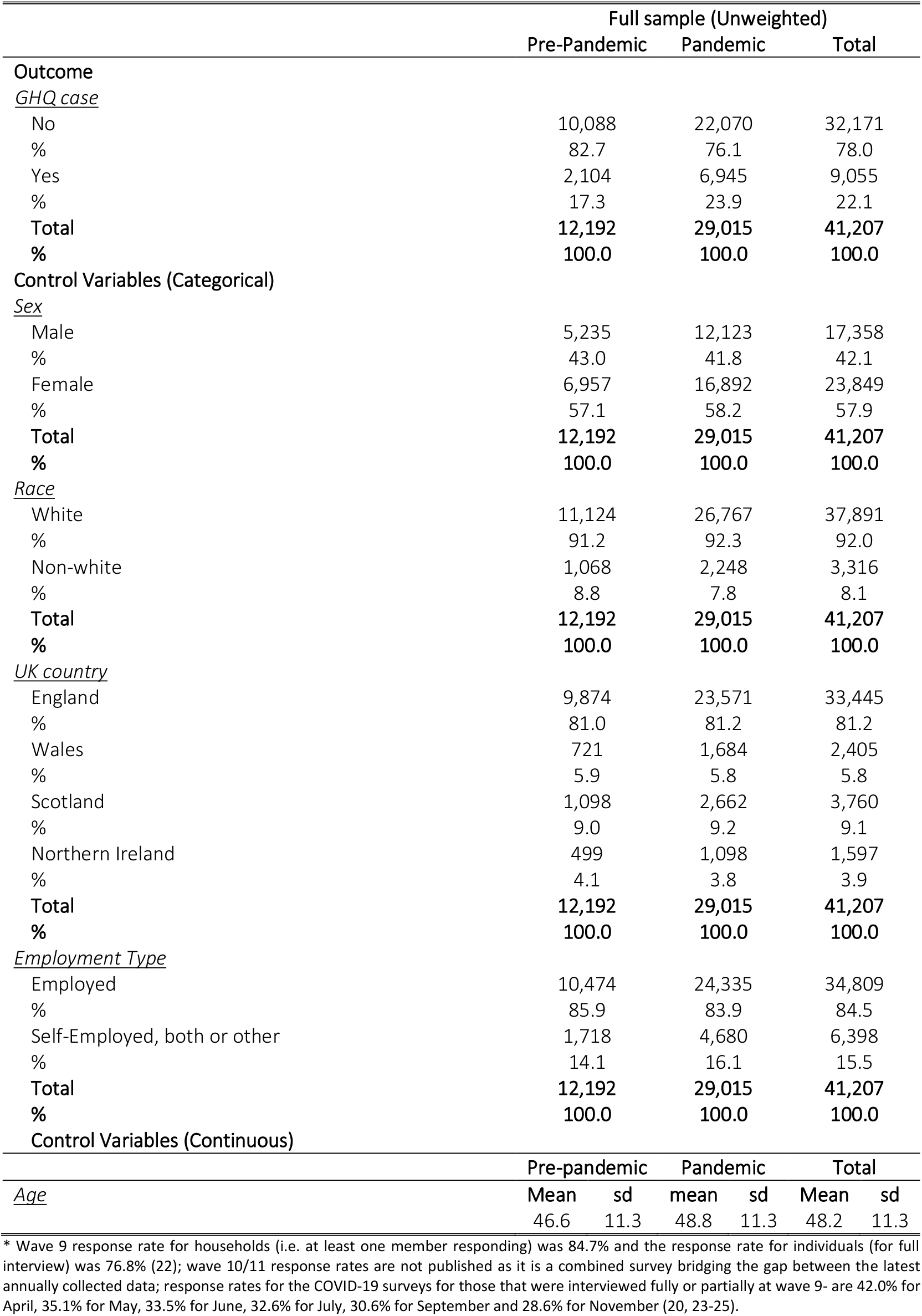
Sample characteristics in the pre and pandemic USoc Waves* (Unweighted Sample)

|  | Full sample (Unweighted) |  |  |  |  |  |
| --- | --- | --- | --- | --- | --- | --- |
|  | Pre-Pandemic |  | Pandemic |  | Total |  |
| Outcome |  |  |  |  |  |  |
| *GHQ case* |  |  |  |  |  |  |
| No | 10,088 |  | 22,070 |  | 32,171 |  |
| % | 82.7 |  | 76.1 |  | 78.0 |  |
| Yes | 2,104 |  | 6,945 |  | 9,055 |  |
| % | 17.3 |  | 23.9 |  | 22.1 |  |
| Total | 12,192 |  | 29,015 |  | 41,207 |  |
| % | 100.0 |  | 100.0 |  | 100.0 |  |
| Control Variables (Categorical) |  |  |  |  |  |  |
| *Sex* |  |  |  |  |  |  |
| Male | 5,235 |  | 12,123 |  | 17,358 |  |
| % | 43.0 |  | 41.8 |  | 42.1 |  |
| Female | 6,957 |  | 16,892 |  | 23,849 |  |
| % | 57.1 |  | 58.2 |  | 57.9 |  |
| Total | 12,192 |  | 29,015 |  | 41,207 |  |
| % | 100.0 |  | 100.0 |  | 100.0 |  |
| *Race* |  |  |  |  |  |  |
| White | 11,124 |  | 26,767 |  | 37,891 |  |
| % | 91.2 |  | 92.3 |  | 92.0 |  |
| Non-white | 1,068 |  | 2,248 |  | 3,316 |  |
| % | 8.8 |  | 7.8 |  | 8.1 |  |
| Total | 12,192 |  | 29,015 |  | 41,207 |  |
| % | 100.0 |  | 100.0 |  | 100.0 |  |
| *UK country* |  |  |  |  |  |  |
| England | 9,874 |  | 23,571 |  | 33,445 |  |
| % | 81.0 |  | 81.2 |  | 81.2 |  |
| Wales | 721 |  | 1,684 |  | 2,405 |  |
| % | 5.9 |  | 5.8 |  | 5.8 |  |
| Scotland | 1,098 |  | 2,662 |  | 3,760 |  |
| % | 9.0 |  | 9.2 |  | 9.1 |  |
| Northern Ireland | 499 |  | 1,098 |  | 1,597 |  |
| % | 4.1 |  | 3.8 |  | 3.9 |  |
| Total | 12,192 |  | 29,015 |  | 41,207 |  |
| % | 100.0 |  | 100.0 |  | 100.0 |  |
| *Employment Type* |  |  |  |  |  |  |
| Employed | 10,474 |  | 24,335 |  | 34,809 |  |
| % | 85.9 |  | 83.9 |  | 84.5 |  |
| Self-Employed, both or other | 1,718 |  | 4,680 |  | 6,398 |  |
| % | 14.1 |  | 16.1 |  | 15.5 |  |
| Total | 12,192 |  | 29,015 |  | 41,207 |  |
| % | 100.0 |  | 100.0 |  | 100.0 |  |
| Control Variables (Continuous) |  |  |  |  |  |  |
|  | Pre-pandemic |  | Pandemic |  | Total |  |
| *Age* | Mean | sd | mean | sd | Mean | sd |
|  | 46.6 | 11.3 | 48.8 | 11.3 | 48.2 | 11.3 |

\* Wave 9 response rate for households (i.e. at least one member responding) was 84.7% and the response rate for individuals (for full interview) was 76.8% (22); wave 10/11 response rates are not published as it is a combined survey bridging the gap between the latest annually collected data; response rates for the COVID-19 surveys for those that were interviewed fully or partially at wave 9- are 42.0% for April, 35.1% for May, 33.5% for June, 32.6% for July, 30.6% for September and 28.6% for November (20, 23-25).

### Trends in GHQ-12 caseness over time, by industry, social class and occupation

Figure 1 shows the weighted prevalence of GHQ-12 caseness by Usoc survey and exposure grouping. In the pre-pandemic period, GHQ-12 caseness remained relatively stable for most exposures. This trend supports combining both surveys as our pre-pandemic period. During the pandemic, GHQ-12 caseness increased considerably across all industries at the beginning (April & May; coinciding with the first UK lockdown -announced 16/03/2020), apart from ‘Agriculture Forestry and Fishing’ (albeit with a small sample size), and across all social class groups, and occupations.

**Figure 1.**
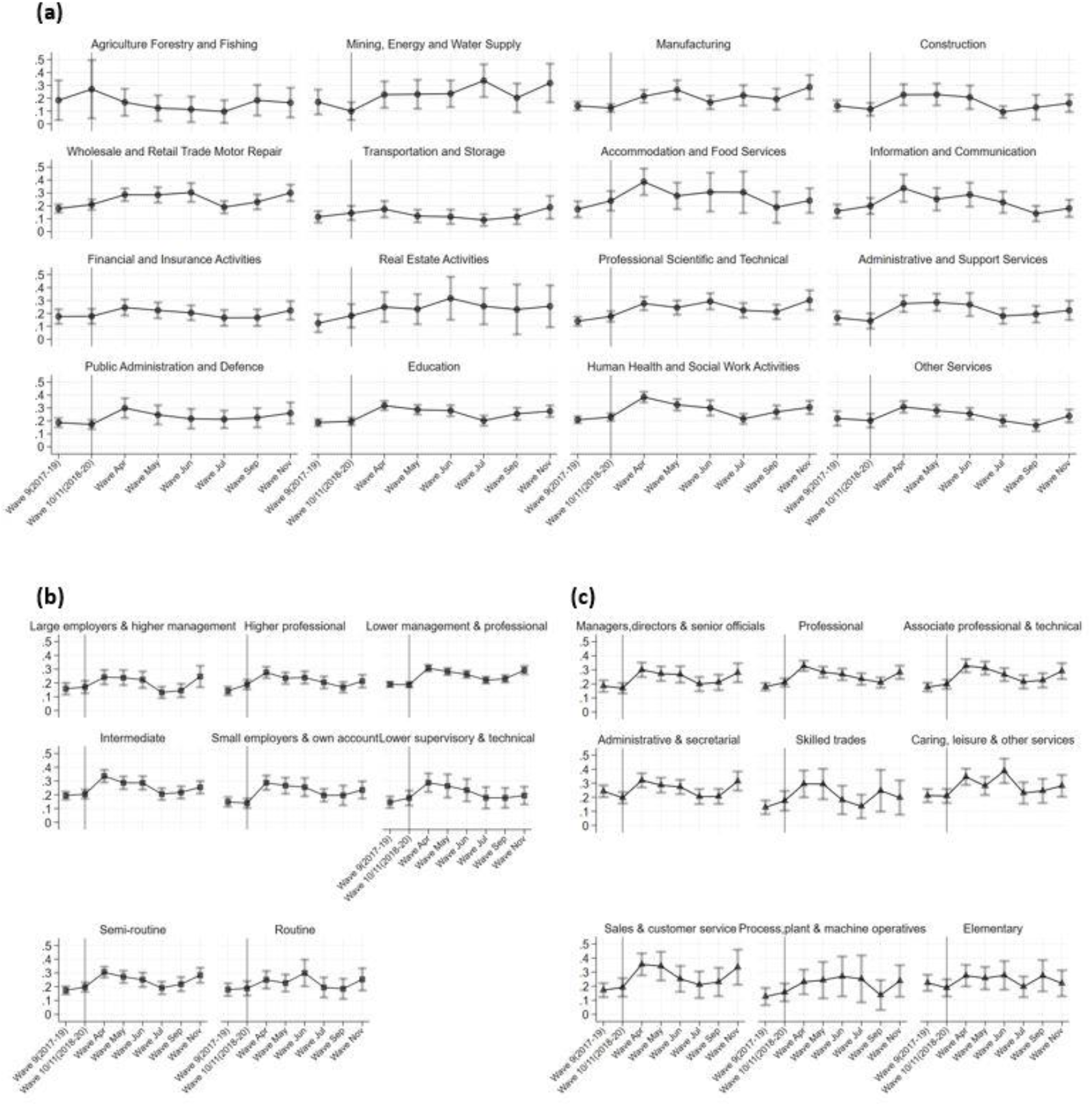
Proportion of GHQ caseness by (a) industry, (b) social class and (c) occupation

GHQ-12 caseness subsequently decreased gradually or remained stable until November when the second lockdown in England was announced (31/10/2020). Prevalence of GHQ-12 caseness in this survey rose to levels similar to the first lockdown period. Comparing the second starting lockdown month (November) to the first (April), most industries displayed similar or higher levels of GHQ-12 caseness. A similar pattern was observed for social class; however, no social class category showed higher prevalence in November compared to April. By occupation, prevalence of GHQ-12 caseness resembled that at the start of the first lockdown, with workers in ‘Skilled trades’ and ‘Elementary’ occupations (e.g., labourers, cleaners, street services workers etc) less affected in the second lockdown.

### Change in GHQ-12 caseness, by industry, social class and occupation

Odds of GHQ-12 caseness almost doubled for most industries in the pandemic period (Table 2); and trebled for the ‘Professional Scientific and Technical’ (OR:3.15, 95% CI 2.17–4.57) and ‘Manufacturing’ (OR:3.01, 95% CI 1.92–4.74) industries. GHQ-12 caseness by social class displayed similar trends, with the likelihood of GHQ-12 caseness in the pandemic period being larger across all categories and ranging from 1.66 (OR:1.66, 95% CI 1.06–2.58) for ‘Routine’ class to more than 3-times (OR:3.24, 95% CI 2.28– 4.63) for ‘Small employers and own account’. Examining exposure by occupation, all groups showed increases in GHQ-12 caseness, with a three-fold increase in odds for ‘Sales and Customer service’ (OR:3.01, 95% CI 1.61–5.62) and a comparable increase for ‘Skilled trades occupations’ (OR:2.88, 95% CI 1.48–5.60). The adjusted results demonstrate increased odds, compared to unadjusted, for all exposures. Sensitivity analysis with a lower GHQ-12 caseness cut-off showed few differences mainly due to categories with low precision (Table S7).

**Table 2:**
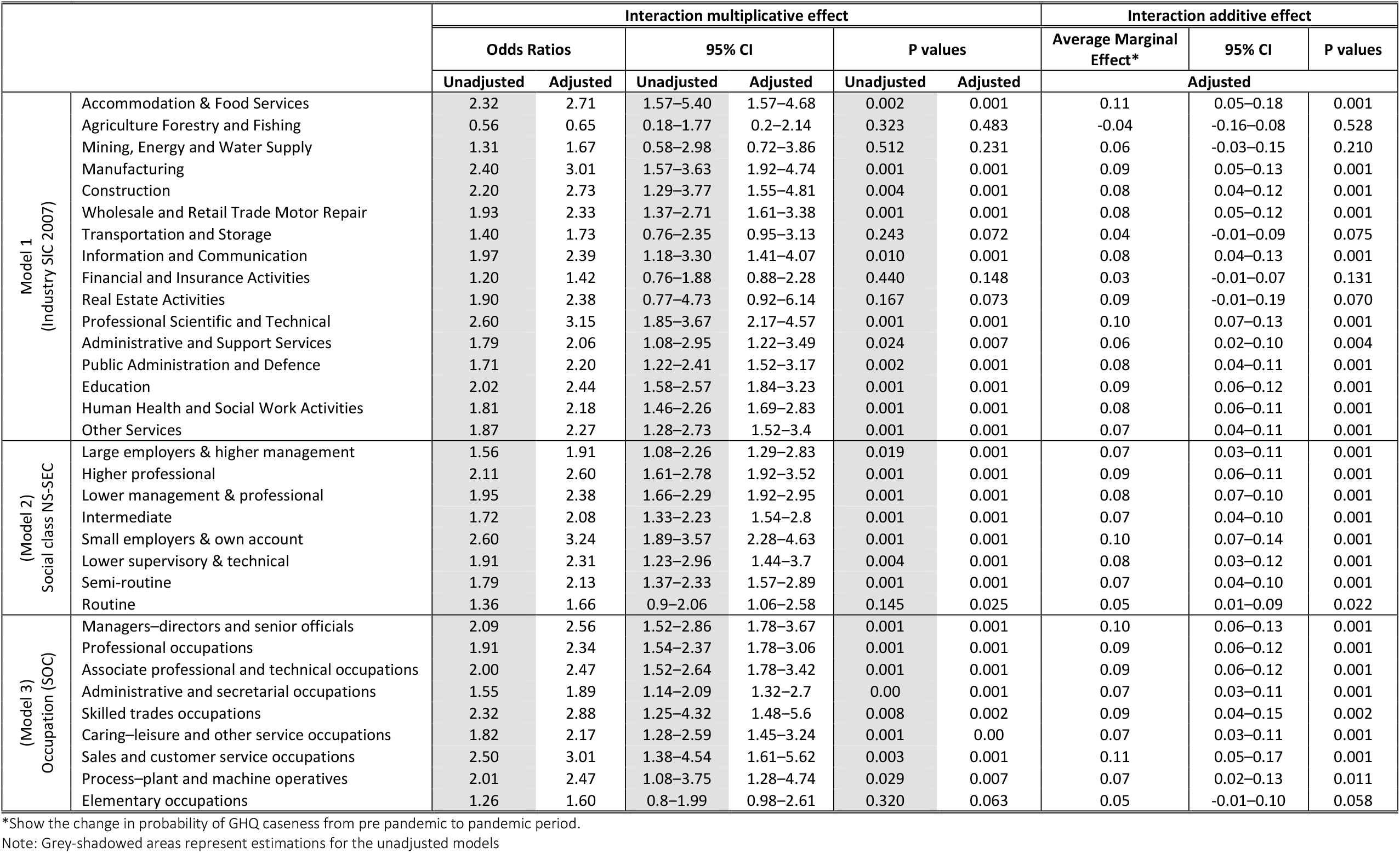
Unstratified Regression models (Models 1, 2 and 3)

|  |  | Interaction multiplicative effect |  |  |  |  |  | Interaction additive effect |  |  |
| --- | --- | --- | --- | --- | --- | --- | --- | --- | --- | --- |
|  |  | Odds Ratios |  | 95% CI |  | P values |  | Average Marginal Effect* | 95% CI | P values |
|  |  | Unadjusted | Adjusted | Unadjusted | Adjusted | Unadjusted | Adjusted | Adjusted |  |  |
| Model 1<br>(Industry SIC 2007) | Accommodation & Food Services | 2.32 | 2.71 | 1.57–5.40 | 1.57–4.68 | 0.002 | 0.001 | 0.11 | 0.05–0.18 | 0.001 |
|  | Agriculture Forestry and Fishing | 0.56 | 0.65 | 0.18–1.77 | 0.2–2.14 | 0.323 | 0.483 | -0.04 | -0.16–0.08 | 0.528 |
|  | Mining, Energy and Water Supply | 1.31 | 1.67 | 0.58–2.98 | 0.72–3.86 | 0.512 | 0.231 | 0.06 | -0.03–0.15 | 0.210 |
|  | Manufacturing | 2.40 | 3.01 | 1.57–3.63 | 1.92–4.74 | 0.001 | 0.001 | 0.09 | 0.05–0.13 | 0.001 |
|  | Construction | 2.20 | 2.73 | 1.29–3.77 | 1.55–4.81 | 0.004 | 0.001 | 0.08 | 0.04–0.12 | 0.001 |
|  | Wholesale and Retail Trade Motor Repair | 1.93 | 2.33 | 1.37–2.71 | 1.61–3.38 | 0.001 | 0.001 | 0.08 | 0.05–0.12 | 0.001 |
|  | Transportation and Storage | 1.40 | 1.73 | 0.76–2.35 | 0.95–3.13 | 0.243 | 0.072 | 0.04 | -0.01–0.09 | 0.075 |
|  | Information and Communication | 1.97 | 2.39 | 1.18–3.30 | 1.41–4.07 | 0.010 | 0.001 | 0.08 | 0.04–0.13 | 0.001 |
|  | Financial and Insurance Activities | 1.20 | 1.42 | 0.76–1.88 | 0.88–2.28 | 0.440 | 0.148 | 0.03 | -0.01–0.07 | 0.131 |
|  | Real Estate Activities | 1.90 | 2.38 | 0.77–4.73 | 0.92–6.14 | 0.167 | 0.073 | 0.09 | -0.01–0.19 | 0.070 |
|  | Professional Scientific and Technical | 2.60 | 3.15 | 1.85–3.67 | 2.17–4.57 | 0.001 | 0.001 | 0.10 | 0.07–0.13 | 0.001 |
|  | Administrative and Support Services | 1.79 | 2.06 | 1.08–2.95 | 1.22–3.49 | 0.024 | 0.007 | 0.06 | 0.02–0.10 | 0.004 |
|  | Public Administration and Defence | 1.71 | 2.20 | 1.22–2.41 | 1.52–3.17 | 0.002 | 0.001 | 0.08 | 0.04–0.11 | 0.001 |
|  | Education | 2.02 | 2.44 | 1.58–2.57 | 1.84–3.23 | 0.001 | 0.001 | 0.09 | 0.06–0.12 | 0.001 |
|  | Human Health and Social Work Activities | 1.81 | 2.18 | 1.46–2.26 | 1.69–2.83 | 0.001 | 0.001 | 0.08 | 0.06–0.11 | 0.001 |
|  | Other Services | 1.87 | 2.27 | 1.28–2.73 | 1.52–3.4 | 0.001 | 0.001 | 0.07 | 0.04–0.11 | 0.001 |
| Model 2)<br>Social class NS-SEC | Large employers & higher management | 1.56 | 1.91 | 1.08–2.26 | 1.29–2.83 | 0.019 | 0.001 | 0.07 | 0.03–0.11 | 0.001 |
|  | Higher professional | 2.11 | 2.60 | 1.61–2.78 | 1.92–3.52 | 0.001 | 0.001 | 0.09 | 0.06–0.11 | 0.001 |
|  | Lower management & professional | 1.95 | 2.38 | 1.66–2.29 | 1.92–2.95 | 0.001 | 0.001 | 0.08 | 0.07–0.10 | 0.001 |
|  | Intermediate | 1.72 | 2.08 | 1.33–2.23 | 1.54–2.8 | 0.001 | 0.001 | 0.07 | 0.04–0.10 | 0.001 |
|  | Small employers & own account | 2.60 | 3.24 | 1.89–3.57 | 2.28–4.63 | 0.001 | 0.001 | 0.10 | 0.07–0.14 | 0.001 |
|  | Lower supervisory & technical | 1.91 | 2.31 | 1.23–2.96 | 1.44–3.7 | 0.004 | 0.001 | 0.08 | 0.03–0.12 | 0.001 |
|  | Semi-routine | 1.79 | 2.13 | 1.37–2.33 | 1.57–2.89 | 0.001 | 0.001 | 0.07 | 0.04–0.10 | 0.001 |
|  | Routine | 1.36 | 1.66 | 0.9–2.06 | 1.06–2.58 | 0.145 | 0.025 | 0.05 | 0.01–0.09 | 0.022 |
| Model 3)<br>Occupation (SOC) | Managers–directors and senior officials | 2.09 | 2.56 | 1.52–2.86 | 1.78–3.67 | 0.001 | 0.001 | 0.10 | 0.06–0.13 | 0.001 |
|  | Professional occupations | 1.91 | 2.34 | 1.54–2.37 | 1.78–3.06 | 0.001 | 0.001 | 0.09 | 0.06–0.12 | 0.001 |
|  | Associate professional and technical occupations | 2.00 | 2.47 | 1.52–2.64 | 1.78–3.42 | 0.001 | 0.001 | 0.09 | 0.06–0.12 | 0.001 |
|  | Administrative and secretarial occupations | 1.55 | 1.89 | 1.14–2.09 | 1.32–2.7 | 0.00 | 0.001 | 0.07 | 0.03–0.11 | 0.001 |
|  | Skilled trades occupations | 2.32 | 2.88 | 1.25–4.32 | 1.48–5.6 | 0.008 | 0.002 | 0.09 | 0.04–0.15 | 0.002 |
|  | Caring–leisure and other service occupations | 1.82 | 2.17 | 1.28–2.59 | 1.45–3.24 | 0.001 | 0.00 | 0.07 | 0.03–0.11 | 0.001 |
|  | Sales and customer service occupations | 2.50 | 3.01 | 1.38–4.54 | 1.61–5.62 | 0.003 | 0.001 | 0.11 | 0.05–0.17 | 0.001 |
|  | Process–plant and machine operatives | 2.01 | 2.47 | 1.08–3.75 | 1.28–4.74 | 0.029 | 0.007 | 0.07 | 0.02–0.13 | 0.011 |
|  | Elementary occupations | 1.26 | 1.60 | 0.8–1.99 | 0.98–2.61 | 0.320 | 0.063 | 0.05 | -0.01–0.10 | 0.058 |

\*Show the change in probability of GHQ caseness from pre pandemic to pandemic period.
Note: Grey-shadowed areas represent estimations for the unadjusted models

Figure 2 shows predicted absolute changes in mental health over the pandemic period (as opposed to relative changes shown by the ORs previously). While the direction of the results remained the same (e.g., ‘Agriculture, Forestry and Fishing’ saw an absolute decline in GHQ-12 caseness while all other industries experienced increases), the industries hardest hit varied. The ‘Professional Scientific and Technical’ and ‘Manufacturing’ industries experienced the biggest relative increase in GHQ-12 caseness, but ‘Accommodation and Food Services’ experienced the greatest absolute increase (11%). All social class groups saw absolute increases in GHQ-12 caseness, and in-line with the ORs results ‘Small employers and own account’ saw the biggest absolute increase (Figure 2b1). Finally, for occupation, ‘Sales and customer service’, ‘Skilled trades’ and ‘Associate professional and technical’ occupations saw large absolute increases in GHQ-12 caseness; however, change among ‘Managers Directors and senior officials’ and ‘Professional’ occupations was comparable.

**Figure 2.**
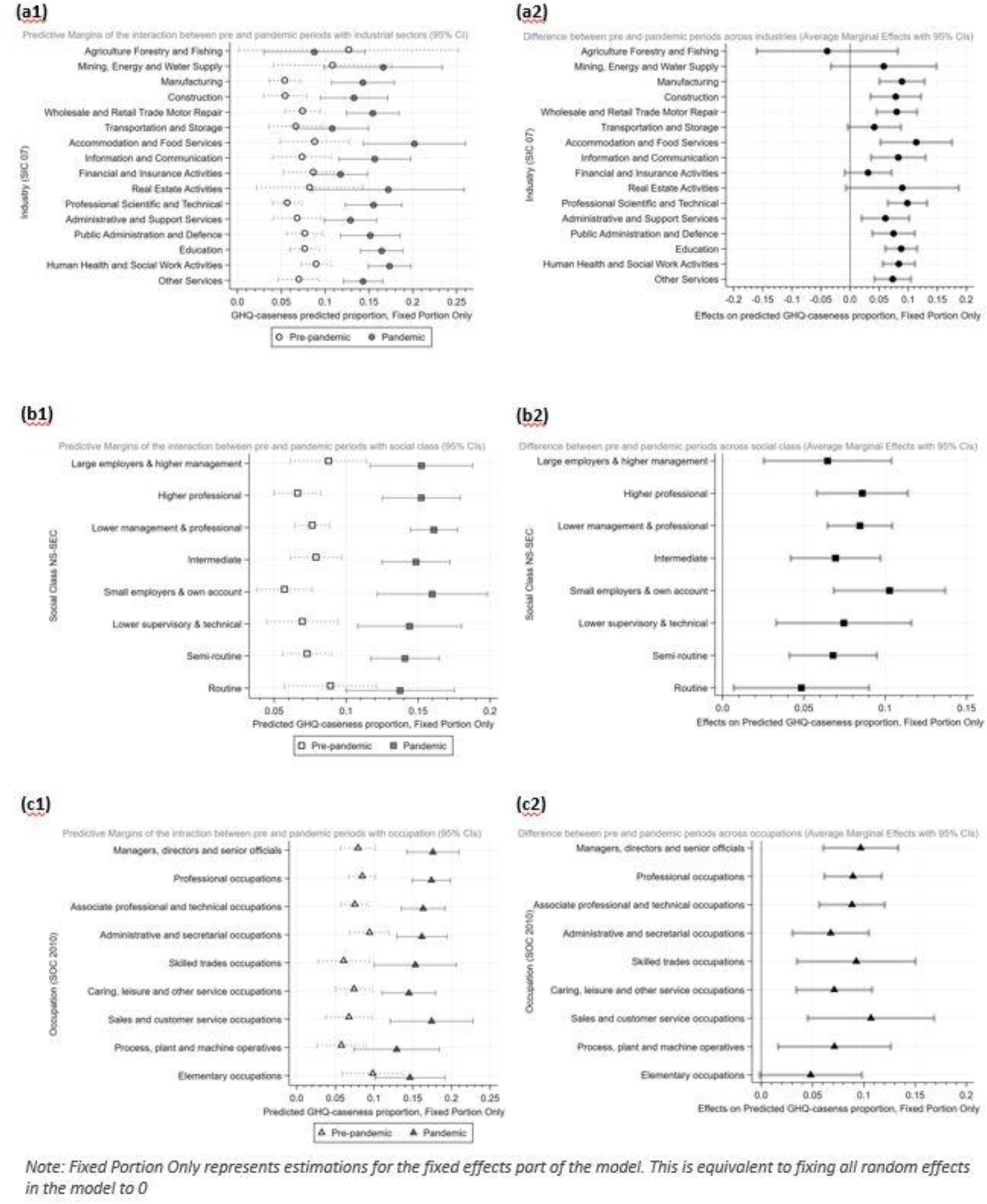
Predictive margins and average marginal effects for all exposures for pre and pandemic periods (Figure 2a1-2c1) and the change in probability during the pandemic period (Figure 2a2-2c2).

### Gender-, age- and UK country of residence- specific differences in GHQ-12 caseness

Analyses where UK country of residence was a part of the 3-way interaction demonstrated little evidence of differences. Similar findings were observed for age. Exceptions were found in the ‘Public Administration and Defence’ and ‘Construction’ industries, where non-England residents and younger workers, respectively, displayed greater differences in GHQ-12 caseness. By social class, older ‘Routine’ and younger ‘Semi-Routine’ workers fared worse, whereas by occupation non-England workers in ‘Elementary’ occupations, and younger ‘professionals’ (Figures S1-S2) displayed differences in GHQ-12 caseness.

Regarding gender, differences in GHQ-12 caseness were apparent in several industries (Figure 3). The effects were consistently larger for women in ‘Transportation and Storage’ (Δ(AVE): −15.9pp; 95% CI: - 28.89; −2.85), ‘Education’ (Δ(AVE): −8.2pp; 95% CI: −13.36; −2.55), and ‘Other Services’ (Δ(AVE): −6.4; 95% CI: −12.18; −0.59). Other industries showed similarly larger effects for women but at a lower CI level (Figure 3). By social class, across most groups mental health deteriorated for both genders, but in ‘Lower management & professional’ (ΔAVE: −7.2pp; 95% CI: −10.34; −4.00) women were affected more. Indication of gender differences for women were observed in other social class groups, albeit at lower CIs level (Figure 3). By occupation, women in ‘Sales & customer service’ exhibited the largest difference (ΔAVE: −13.4pp; 95% CI: −24.47; −2.29) followed by ‘Professional occupations’ (ΔAVE: −5.0pp; 95% CI: - 9.74; −0.29). Similar to industry and social class, differences were also observed in other occupations at a lower CI level (Figure 3).

**Figure 3.**
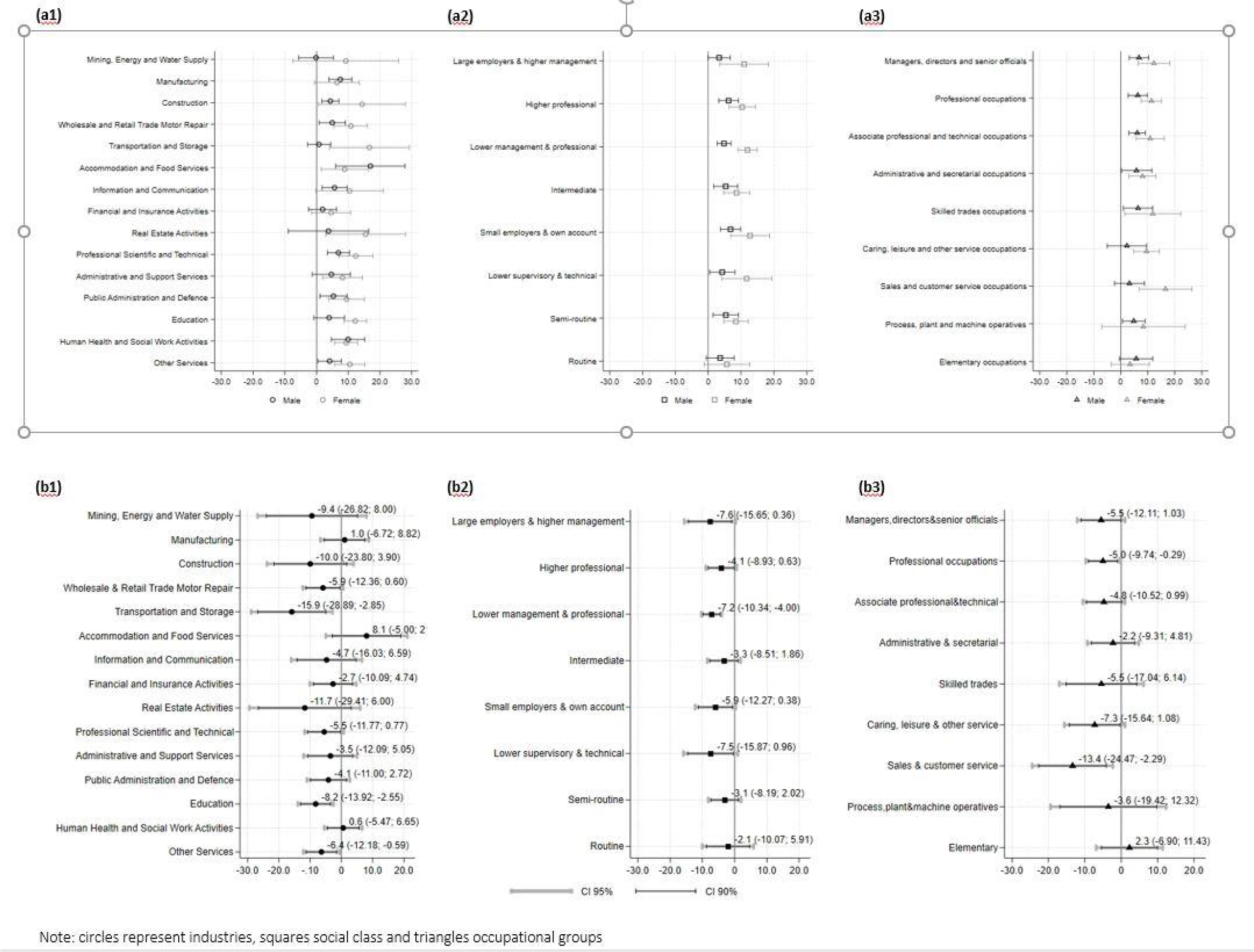
Average Marginal Effects (AVE) by gender for all exposures (a1,a2,a3) and their percentage point probability difference (Δ(AVE) (b1,b2,b3)

## Discussion

### Summary of findings

Psychological distress increased substantially in UK workers during the COVID-19 pandemic across almost all industries, social class and occupation exposure groups. In most industries the odds of GHQ-12 caseness increased more than twofold, with workers in ‘professional, scientific, technical’ and the ‘hospitality’, ‘construction’ and ‘manufacturin’g industries being most impacted. By social class, small employers and the self-employed were most adversely affected, with odds increasing more than 3 times, while for most of the other social class groups the odds of GHQ-12 caseness increased more than two-fold. During the pandemic, occupations in ‘sales, customer service’ and ‘skilled trades’ were most affected. Analysis using 3-way interactions, showed little evidence of significant differences in odds of GHQ-12 caseness by UK country of residence and age (with a few exceptions). Marked differences for women were observed in all exposure groups and particularly in the ‘Transportation & Storage’ and ‘Education’ industries, the ‘Lower supervisory & Technical’ social class group and in ‘Sales & customer service’ occupations.

### Findings in context with previous literature

Our findings are in line with national statistics demonstrating that rates of work-related stress, depression and anxiety have been increasing (30). Estimates from the Labour Force Survey show that the prevalence of work-related stress, depression or anxiety in 2019/20 was substantially higher than the previous period (30). The UK’s Health & Safety Executive reports that stress, depression or anxiety is more prevalent in public service industries, such as education; health and social care; and public administration and defence. While our findings support this, we also show the impact of the pandemic on a number of other industries (e.g., ‘Professional Scientific and Technical’, ‘Accommodation and Food Services’, ‘Manufacturing’ and ‘Construction’). By occupation, professional occupations that are common across public service industries (such as healthcare workers; teaching professionals and public service professionals) showed higher levels of stress compared to all jobs (30). Similarly, our findings demonstrated an increase in GHQ-12 caseness across most occupations, however workers in some occupations with high levels of unstable working conditions, low autonomy and job control, technical tasks, and female workers in occupations such as sales and customer service representatives and professionals were most adversely affected.

Most research to date during the pandemic has focused on the health and wellbeing of healthcare workers, where significantly higher levels of stress, burnout, secondary trauma, anxiety, and depression have been recorded among professionals working with COVID-19 patients (31). Other frontline workers, working in areas with higher rates of contagion also report higher levels of stress and burnout and lower levels of compassion satisfaction (31). Our results support the findings that frontline healthcare workers require further support, and that targeted prevention and intervention programs are necessary. However, we also demonstrate that other industries and occupational groups are at risk and require specific attention. These may not have the stability or safety nets in place and/or could not continue with their occupations by working from home, including those in manufacturing, construction, hospitality, the self-employed and small business owners, and occupations that involve unstable working conditions or the performance of technical tasks.

### Study strengths and limitations

Our study has several strengths. We used a large nationally representative longitudinal dataset to examine differences in psychological distress during the COVID-19 pandemic across industries, social class groups, and occupations, which fills an important gap in the literature. Our analysis included pre-pandemic outcome measures and six surveys of data collection after the start of the pandemic, and therefore we were able to examine trends before and after the initial lockdown. We also explored multiple dimensions of employment and explored heterogeneity across population groups. Some limitations should be noted. First, while estimates were weighted to adjust for survey non-participation there may still have been some residual bias, as response rates in the COVID surveys were lower than usual. Second, there were changes in the modality of administration of the COVID-19 surveys compared to pre-pandemic surveys (from mixed mode: face-to-face/web/phone, to online), which may have made modest contributions to the changes reported. However, empirical investigation suggests this is unlikely to have biased responses (32). The pandemic context may have also influenced participant reporting more broadly. Furthermore, we need to be cautious as observations in some industries (Agriculture, Forestry and Fishing: N=342; Mining, Energy and Water Supply: N=744 and Real Estate Activities: N=485) were much lower than in others reducing the precision of estimates. The stratified analysis displayed relatively small differences and wide confidence intervals and we suggest these analyses be replicated when data from more COVID-19 surveys are available. Discrepancies between social class and occupational exposure groups that may cover the same jobs (e.g., ‘Routine’ social class and ‘Elementary’ occupations) highlight the need to use more detailed occupational breakdowns as some specific occupations may have unique and greater risks for poor mental health, such as greater risk of infection and/or financial hardship. We also only included one measure of mental health and results may differ using other self-reported measures of depression or anxiety. Further research examining clinical outcomes, such as receipt of antidepressant prescription, is also needed.

### Implications for policy and practice

Our study has important implications for occupational health and public health policy. The increase in psychological distress seen in UK workers highlights the trade-off and problem shifting required to protect the public’s physical health during the pandemic and the adverse effects this could have. Our findings of substantial disparities in GHQ-12 caseness across industries, social class groups, occupations, as well as some differences between genders and regions, highlight the complexity of the extended health harms associated with the pandemic and risks to mental health. These can include uncertainty about career prospects and the unpredictability of the labour market, fear of job and/or income loss, and worry about financial hardship or increased job demands. Concurrently, worries about exposure risks to the virus while at work will also differ across sectors and occupations. Finding that particular groups have been disproportionately affected illustrates broader inequalities in the job market and employment conditions (11 12 21 33). The differences by UK country of residence may reflect pre-existing labour markets, as well as de-centralised handling of the lockdown measures. The self-employed and small business owners may have fewer safety nets and greater fear of the lack of employment opportunities after the pandemic. Further research is needed to understand the risk factors and mechanisms that are driving these findings for each group and whether the substantial increase in psychological distress remains as lockdown measures are eased and working environments and conditions establish their new ‘normal’. The deterioration of mental health is also expected to create additional capacity problems for primary and secondary care services in the UK and internationally, given the strain these services are already under due to the COVID-19 pandemic. Monitoring the mental health of the working-age population can inform workplace and non-workplace support measures that are needed.

## Concluding Remarks

The COVID-19 pandemic has substantially affected the mental health of the working-age population. However, certain industries, social class groups, occupations, and socio-demographic groups were disproportionately affected. The working environment under the COVID-19 pandemic as well as the anticipated ‘new normal’ in employment opportunities, patterns and conditions that may follow, are likely to include a degree of fear, uncertainty, employment instability and job and/or income loss. Employers need to consider tailored workplace practices for mental health problems. Furthermore, policies tailored to these exposures are needed prioritising for instance support to the self-employed/small business owners, and particular demographic groups (e.g., women in sales and customer service occupations, younger ‘construction’ or non-England workers in ‘Public Administration and Defence’) with high risk.

## Data Availability

All data are available online from Understanding Society (https://www.understandingsociety.ac.uk/)

## Acknowledgements

We thank all Understanding Society participants.

## Author contributions

ED conceived the idea for the study. TK, MG, SVK and ED designed the study. TK led and conducted the statistical analysis. TK and ED drafted the manuscript. All authors contributed to the interpretation of the results, critically revised the paper and agreed on the final version for submission.

