## Supplementary material for "Population-level changes in the mental health of UK workers during the COVID-19 pandemic: A longitudinal study using Understanding Society"

Table S1A. SIC Industrial Classifications Sections in Understanding Society and study

| SIC Industrial Classifications Sections - Understanding Society | | SIC Industrial Classifications Sections -Included in Study |
|---|---|---|
| Agriculture, Forestry and Fishing | 1 | Agriculture, Forestry and Fishing |
| Mining and Quarrying | 2 | Mining, Energy and Water Supply |
| Electricity, Gas, Steam and Air Conditioning | | |
| Water Supply; Sewerage, Waste Management and Remediation Activities | | |
| Manufacturing | 3 | Manufacturing |
| Construction | 4 | Construction |
| Wholesale and Retail Trade | 5 | Wholesale and Retail Trade Motor Repair |
| Repair of Motor Vehicles and Motorcycles | | |
| Transportation and Storage | 6 | Transportation and Storage |
| Accommodation and Food Service Activities | 7 | Accommodation and Food Services |
| Information and Communication | 8 | Information and Communication |
| Financial and Insurance Activities | 9 | Financial and Insurance Activities |
| Real Estate Activities | 10 | Real Estate Activities |
| Professional, Scientific and Technical Activities | 11 | Professional Scientific and Technical |
| Administrative and Support Service Activities | 12 | Administrative and Support Services |
| Public Administration and Defence; Compulsory Social Security | 13 | Public Administration and Defence |
| Education | 14 | Education |
| Human Health and Social Work Activities | 15 | Human Health and Social Work Activities |
| Arts, Entertainment and Recreation | 16 | Other Services |
| Other Service Activities | | |
| Activities of Households as Employers | | |

Table S1B. Social class by occupation

| | Managers, directors and senior officials | Professional occupations | Associate professional and technical occupations | Administrative and secretarial occupations | Skilled trades occupations | Caring, leisure and other service occupations | Sales and customer service occupations | Process, plant and machine operatives | Elementary occupations | Total |
|---|---|---|---|---|---|---|---|---|---|---|
| Large employers & higher management | 1,026 | 371 | 280 | 26 | 14 | 0 | 15 | 6 | 2 | 1,740 |
| % | 30.81 | 5.43 | 5.32 | 0.74 | 1.03 | 0 | 0.97 | 0.53 | 0.12 | 6.42 |
| Higher professional | 60 | 2,444 | 226 | 28 | 14 | 11 | 10 | 7 | 0 | 2,800 |
| % | 1.80 | 35.79 | 4.29 | 0.79 | 1.03 | 0.45 | 0.65 | 0.61 | 0 | 10.33 |
| Lower management & professional | 1,799 | 3,498 | 3,722 | 795 | 146 | 117 | 203 | 34 | 47 | 10,361 |
| % | 54.02 | 51.22 | 70.69 | 22.48 | 10.75 | 4.79 | 13.11 | 2.98 | 2.84 | 38.22 |
| Intermediate | 59 | 271 | 601 | 2,030 | 90 | 395 | 362 | 11 | 21 | 3,840 |
| % | 1.77 | 3.97 | 11.42 | 57.39 | 6.63 | 16.16 | 23.37 | 0.96 | 1.27 | 14.16 |
| Small employers & own account | 309 | 195 | 160 | 187 | 393 | 315 | 80 | 152 | 177 | 1,968 |
| % | 9.28 | 2.86 | 3.04 | 5.29 | 28.94 | 12.89 | 5.16 | 13.31 | 10.69 | 7.26 |
| Lower supervisory & technical | 46 | 5 | 110 | 58 | 426 | 173 | 113 | 274 | 169 | 1,374 |
| % | 1.38 | 0.07 | 2.09 | 1.64 | 31.37 | 7.08 | 7.3 | 23.99 | 10.21 | 5.07 |
| Semi-routine | 30 | 39 | 146 | 381 | 184 | 1,334 | 748 | 243 | 509 | 3,614 |
| % | 0.90 | 0.57 | 2.77 | 10.77 | 13.55 | 54.58 | 48.29 | 21.28 | 30.74 | 13.33 |
| Routine | 1 | 6 | 20 | 32 | 91 | 99 | 18 | 415 | 731 | 1,413 |
| | 0.03 | 0.09 | 0.38 | 0.9 | 6.7 | 4.05 | 1.16 | 36.34 | 44.14 | 5.21 |
| Total | **3,330** | **6,829** | **5,265** | **3,537** | **1,358** | **2,444** | **1,549** | **1,142** | **1,656** | **27,110** |
| | **100** | **100** | **100** | **100** | **100** | **100** | **100** | **100** | **100** | **100** |

### Weighting strategy

First, we created an inverse-probability weight within each survey wave for having outcome data based on:

i. Age, split in three broad age bands (1: 18/29y old, 2: 30/45y old and 3: Over 45y old),
ii. gender (male vs female),
iii. race (white vs non-white) and
iv. type of employment which is another binary variable that categorises employees versus those that are either self-employed, both employees and self-employed and other.

These were combined with weights provided with the COVID-19 surveys, which were designed to weight the observed sample at each COVID survey to resemble the sample from the 2017-2019 survey. We also weighted the 2018-2020 survey to resemble the 2017-2019 survey based on the same covariates listed above plus the binary variable for psychological distress and combined this with the weight for outcome missingness for that survey. By using this method, each individual has a non-monotone weight at each survey wave.

Table S2.: Transition probabilities and correlations between current and lagged values for industry, social class, and occupation

| Industry | Pre pandemic | | Pandemic | Social class | Pre pandemic | | Occupation | Pre pandemic | |
|---|---|---|---|---|---|---|---|---|---|
| | 2-wave | 1-wave | 1-month PP | | 2-wave | 1-wave | | 2-wave | 1-wave |
| Agriculture Forestry and Fishing | 93.74 | 94.02 | 90.32 | Large employers & higher management | 85.59 | 85.64 | Managers, directors and senior officials | 87.29 | 87.57 |
| Mining, Energy and Water Supply | 91.91 | 91.79 | 79.51 | Higher professional | 92.83 | 92.72 | Professional occupations | 93.20 | 93.05 |
| Manufacturing | 93.61 | 93.41 | 87.39 | Lower management & professional | 91.85 | 91.79 | Associate professional and technical occupations | 86.19 | 86.29 |
| Construction | 93.20 | 93.55 | 89.69 | Intermediate | 87.38 | 87.26 | Administrative and secretarial occupations | 86.04 | 85.74 |
| Wholesale and Retail Trade Motor Repair | 91.23 | 91.15 | 89.51 | Small employers & own account | 92.23 | 92.24 | Skilled trades occupations | 87.73 | 87.80 |
| Transportation and Storage | 93.95 | 93.94 | 79.52 | Lower supervisory & technical | 83.40 | 83.44 | Caring, leisure and other service occupations | 89.58 | 89.66 |
| Accommodation and Food Services | 84.48 | 84.41 | 74.12 | Semi-routine | 87.22 | 87.19 | Sales and customer service occupations | 80.22 | 80.87 |
| Information and Communication | 92.66 | 92.64 | 79.30 | Routine | 85.65 | 85.79 | Process, plant and machine operatives | 88.95 | 88.84 |
| Financial and Insurance Activities | 93.78 | 93.68 | 90.22 | | | | Elementary occupations | 81.94 | 82.66 |
| Real Estate Activities | 92.69 | 92.67 | 85.51 | | | | | | |
| Professional Scientific and Technical | 92.90 | 92.88 | 72.31 | | | | | | |
| Administrative and Support Services | 87.26 | 87.27 | 54.38 | | | | | | |
| Public Administration and Defence | 94.58 | 94.79 | 77.32 | | | | | | |
| Education | 96.00 | 95.87 | 96.41 | | | | | | |
| Human Health and Social Work Activities | 96.54 | 96.48 | 89.47 | | | | | | |
| Other Services | 90.30 | 90.14 | 62.96 | | | | | | |
| Average | 92.43 | 92.42 | 81.12 | | 88.27 | 88.26 | | 86.79 | 86.94 |

Table S3: Sample characteristics in the pre and pandemic Waves (Weighted Sample)

| | Full sample (Weighted) | | |
|---|---|---|---|
| | Pre-Pandemic | Pandemic | Total |
| **Outcome** | | | |
| *GHQ case %* | | | |
| No | 82.06 | 75.34 | 77.34 |
| Yes | 17.94 | 24.66 | 22.66 |
| **Total** | **100** | **100** | **100** |
| **Control Variables (Categorical)** | | | |
| *Sex %* | | | |
| Male | 46.68 | 49.60 | 48.73 |
| Female | 53.32 | 50.22 | 51.27 |
| **Total** | **100** | **100** | **100** |
| *Race %* | | | |
| White | 93.77 | 93.73 | 93.74 |
| Non-White | 6.23 | 6.27 | 6.26 |
| **Total** | **100** | **100** | **100** |
| *UK country %* | | | |
| England | 86.60 | 85.21 | 85.63 |
| Wales | 3.94 | 4.32 | 4.21 |
| Scotland | 7.81 | 8.06 | 7.98 |
| Northern Ireland | 1.64 | 2.41 | 2.18 |
| **Total** | **100** | **100** | **100** |
| *Employment Type %* | | | |
| Employed | 86.42 | 85.02 | 85.41 |
| Self-Employed, both or other | 13.58 | 14.98 | 14.56 |
| **Total** | **100** | **100** | **100** |

**Control Variables (Continuous)**

| | Pre-pandemic | | Pandemic | | Total | |
|---|---|---|---|---|---|---|
| *Age* | **mean** | **sd** | **mean** | **sd** | **mean** | **sd** |
| | 45.20 | 11.86 | 45.92 | 12.21 | 45.70 | 12.11 |

Table S4: Observations for Industry (SIC-2007) in pre-pandemic and pandemic Waves (Unweighted and Weighted Sample (in brackets))

| Industry SIC-2007 | Full sample | | |
|---|---|---|---|
| | Pre-pandemic | Pandemic | Total |
| Agriculture Forestry and Fishing | 58 | 284 | 342 |
| % | 0.47 (0.45) | 0.98 (1.22) | 0.83 (0.99) |
| Mining, Energy and Water Supply | 171 | 573 | 744 |
| % | 1.4 (1.31) | 1.97 (1.95) | 1.8 (1.76) |
| Manufacturing | 1,038 | 1,877 | 2,915 |
| % | 8.51 (8.54) | 6.47 (7.01) | 7.07 (7.47) |
| Construction | 533 | 1,311 | 1,844 |
| % | 4.37 (4.79) | 4.52 (5.90) | 4.47 (5.57) |
| Wholesale and Retail Trade Motor Repair | 1,326 | 2,748 | 4,074 |
| % | 10.88 (11.86) | 9.47 (11.94) | 9.89 (11.91) |
| Transportation and Storage | 492 | 962 | 1,454 |
| % | 4.04 (4.18) | 3.32 (4.19) | 3.53 (4.19) |
| Accommodation and Food Services | 344 | 670 | 1,014 |
| % | 2.82 (2.93) | 2.31 (2.72) | 2.46 (2.78) |
| Information and Communication | 495 | 1,314 | 1,809 |
| % | 4.06 (4.21) | 4.53 (4.44) | 4.39 (4.37) |
| Financial and Insurance Activities | 430 | 1 354 | 1,784 |
| % | 3.52 (3.86) | 4.67 (4.25) | 4.33 (4.13) |
| Real Estate Activities | 159 | 326 | 485 |
| % | 1.30 (1.32) | 1.12 (0.90) | 1.18 (1.03) |
| Professional Scientific and Technical | 948 | 1 853 | 2,801 |
| % | 7.78 (7.84) | 6.39 (5.37) | 6.80 (6.11) |
| Administrative and Support Services | 465 | 1,292 | 1,757 |
| % | 3.81 (4.02) | 4.45 (3.72) | 4.26 (3.81) |
| Public Administration and Defence | 1,054 | 1,465 | 2,519 |
| % | 8.65 (7.89) | 5.05 (4.51) | 6.11 (5.52) |
| Education | 1,820 | 4,653 | 6,473 |
| % | 14.93 (14.41) | 16.04 (13.59) | 15.71 (13.83) |
| Human Health and Social Work Activities | 2,266 | 4,360 | 6,626 |
| % | 18.59 (17.41) | 15.03 (13.63) | 16.08 (14.76) |
| Other Services | 593 | 3,973 | 4,566 |
| % | 4.86 (4.97) | 13.69 (14.66) | 11.08 (11.77) |
| Total | 12,192 | 29,015 | 41,207 |
| % | 100 | 100 | 100 |

Table S5:Observations for Social Class (NS-SEC) pre-pandemic and pandemic Waves (Unweighted and Weighted Sample (in brackets))

| | Full sample | | |
|---|---|---|---|
| Social Class NS-SEC | **Pre-pandemic** | **Pandemic** | **Total** |
| Large employers & higher management | 715 | 1,686 | 2,401 |
| % | 5.86 (5.92) | 5.81 (5.32) | 5.83 (5.50) |
| Higher professionals | 1,351 | 3,214 | 4,565 |
| % | 11.08 (10.41) | 11.08 (9.15) | 11.08 (9.52) |
| Lower management & pr | 4,370 | 10,455 | 14,825 |
| % | 35.84 (35.17) | 36.03 (31.28) | 35.98 (32.44) |
| Intermediate | 1,697 | 4,164 | 5,861 |
| % | 13.92 (13.99) | 14.35 (13.98) | 14.22 (13.98) |
| Small employers & own account | 1,108 | 2,471 | 3,579 |
| % | 9.09 (8.90) | 8.52 (8.47) | 8.69 (8.60) |
| Lower supervisory & technical | 700 | 1,612 | 2,312 |
| % | 5.74 (6.17) | 5.56 (7.48) | 5.61 (7.09) |
| Semi-routine | 1,622 | 3,888 | 5,510 |
| % | 13.30 (13.88) | 13.4 (16.45) | 13.37 (15.68) |
| Routine | 629 | 1,525 | 2,154 |
| % | 5.16 (5.56) | 5.26 (7.88) | 5.23 (7.19) |
| Total | **12,192** | **29,015** | **41,207** |
| % | **100** | **100** | **100** |

Table S6:Observations for Occupations (SoC) in pre-pandemic and pandemic Waves (Unweighted and Weighted Sample (in brackets))

| Occupations SOC 2010, condensed | Pre-pandemic | Full sample<br>Pandemic | Total |
|---|---|---|---|
| Managers, directors and senior official | 978 | 2,352 | 3,330 |
| % | 11.91 (12.17) | 12.44 (12.01) | 12.28 (12.06) |
| Professional occupations | 2,051 | 4,778 | 6,829 |
| % | 24.97 (23.29) | 25.28 (20.12) | 25.19 (21.07) |
| Associate professional and technical occupations | 1,565 | 3,700 | 5,265 |
| % | 19.06 (18.76) | 19.58 (18.53) | 19.42 (18.60) |
| Administrative and secretarial occupations | 1,055 | 2,482 | 3,537 |
| % | 12.85 (12.51) | 13.13 (10.51) | 13.05 (11.11) |
| Skilled trades occupations | 420 | 938 | 1,358 |
| % | 5.12 (5.72) | 4.96 (7.89) | 5.01 (7.24) |
| Caring leisure and other service occupations | 775 | 1,669 | 2,444 |
| % | 9.44 (9.20) | 8.83 (9.62) | 9.02 (9.49) |
| Sales and customer service occupations | 500 | 1,049 | 1,549 |
| % | 6.09 (6.67) | 5.55 (7.26) | 5.71 (7.09) |
| Process, plant, and machine operatives | 358 | 784 | 1,142 |
| % | 4.36 (4.47) | 4.15 (5.31) | 4.21 (5.06) |
| Elementary occupations | 508 | 1,148 | 1,656 |
| % | 6.19 (7.20) | 6.07 (8.75) | 6.11 (8.28) |
| **Total** | **8,210** | **18,900** | **27,110** |
| **%** | **100** | **100** | **100** |

Table S7. Unstratified Regression models – Sensitivity analysis for outcome variable ((0: 0≤GHQ-12≤2, 1: 3≤GHQ-12≤12).

| | | Odds Ratios | | P values | | CI 95% | |
|---|---|---|---|---|---|---|---|
| | | Analytical Model | Sensitivity | Analytical Model | Sensitivity | Analytical Model | Sensitivity |
| Industry SIC 2007 (Model 1) | Accomm&Food Services | 2.710 | 2.372 | 0.000 | 0.001 | 1.569–4.680 | 1.435–3.939 |
| | Agriculture Forestry and Fishing | 0.655 | 0.525 | 0.483 | 0.301 | 0.201–2.136 | 0.155–1.784 |
| | Mining–Energy and Water Supply | 1.670 | 1.841 | 0.231 | 0.136 | 0.722–3.863 | 0.827–4.117 |
| | Manufacturing | 3.013 | 3.043 | 0.000 | 0.000 | 1.917–4.737 | 2.051–4.531 |
| | Construction | 2.726 | 2.153 | 0.001 | 0.016 | 1.545–4.807 | 1.156–4.031 |
| | Wholesale and Retail Trade Motor Repair | 2.333 | 2.358 | 0.000 | 0.000 | 1.611–3.380 | 1.651–3.373 |
| | Transportation and Storage | 1.725 | 2.014 | 0.072 | 0.014 | 0.952–3.126 | 1.118–3.37 |
| | Information and Communication | 2.393 | 2.107 | 0.001 | 0.001 | 1.406–4.073 | 1.34–3.356 |
| | Financial and Insurance Activities | 1.420 | 1.531 | 0.148 | 0.065 | 0.883–2.282 | 0.977–2.413 |
| | Real Estate Activities | 2.378 | 2.502 | 0.073 | 0.063 | 0.922–6.135 | 0.954–6.596 |
| | Professional Scientific and Technical | 3.150 | 3.461 | 0.000 | 0.000 | 2.169–4.575 | 2.454–4.908 |
| | Administrative and Support Services | 2.061 | 1.852 | 0.007 | 0.019 | 1.218–3.488 | 1.105–3.097 |
| | Public Administration and Defence | 2.197 | 2.356 | 0.000 | 0.000 | 1.522–3.172 | 1.691–3.287 |
| | Education | 2.435 | 2.689 | 0.000 | 0.000 | 1.835–3.232 | 2.047–3.532 |
| | Human Health and Social Work Activities | 2.185 | 2.266 | 0.000 | 0.000 | 1.687–2.830 | 1.788–2.876 |
| | Other Services | 2.273 | 2.155 | 0.000 | 0.000 | 1.517–3.404 | 1.494–3.121 |
| Social class NS-SEC (Model 2) | Large employers & higher management) | 1.907 | 1.893 | 0.001 | 0.001 | 1.287–2.827 | 1.324–2.75 |
| | Higher professional | 2.601 | 2.810 | 0.000 | 0.000 | 1.919–3.524 | 2.111–3.694 |
| | Lower management & professional | 2.381 | 2.477 | 0.000 | 0.000 | 1.924–2.947 | 2.028–3.02 |
| | Intermediate | 2.078 | 2.159 | 0.000 | 0.000 | 1.544–2.796 | 1.622–2.873 |
| | Small employers & own account | 3.244 | 3.180 | 0.000 | 0.000 | 2.276–4.626 | 2.217–4.582 |
| | Lower supervisory & technical | 2.308 | 2.160 | 0.000 | 0.001 | 1.441–3.697 | 1.402–3.345 |
| | Semi-routine | 2.132 | 2.191 | 0.000 | 0.000 | 1.575–2.887 | 1.649–2.914 |
| | Routine | 1.656 | 1.649 | 0.025 | 0.021 | 1.065–2.577 | 1.08–2.531 |
| Occupation (SOC) (Model 3) | Managers, directors and senior officials | 2.557 | 2.369 | 0.000 | 0.000 | 1.783–3.667 | 1.704–3.279 |
| | Professional occupations | 2.337 | 2.621 | 0.000 | 0.000 | 1.783–3.064 | 2.037–3.386 |
| | Associate professional and technical occupations | 2.471 | 2.664 | 0.000 | 0.000 | 1.784–3.424 | 1.954–3.622 |
| | Administrative and secretarial occupations | 1.892 | 1.925 | 0.000 | 0.000 | 1.324–2.703 | 1.382–2.688 |
| | Skilled trades occupations | 2.877 | 3.068 | 0.002 | 0.001 | 1.479–5.597 | 1.571–6.007 |
| | Caring, leisure and other service occupations | 2.166 | 2.364 | 0.000 | 0.000 | 1.448–3.239 | 1.624–3.452 |
| | Sales and customer service occupations | 3.010 | 3.064 | 0.001 | 0.000 | 1.613–5.618 | 1.643–5.647 |
| | Process, plant and machine operatives | 2.467 | 2.457 | 0.007 | 0.003 | 1.283–4.742 | 1.361–4.459 |
| | Elementary occupations | 1.596 | 1.471 | 0.063 | 0.101 | 0.976–2.612 | 0.928–2.337 |

Note: Grey-shadowed areas represent estimations for the sensitivity analysis

Figure S1: Flowchart of study participants

Usoc observations (n) and participants (N) at baseline (Waves 9, 10/11 and all Covid-19 surveys:

| | Pre-Pandemic | Pandemic | Total |
|---|---|---|---|
| n | 69,437 | 86,469 | **155,906** |
| N | | | **40,245** |

Eligible n and N who were interviewed fully or partially:

| | Pre-Pandemic | Pandemic | Total |
|---|---|---|---|
| n | 67,649 | 85,360 | **153,009** |
| N | | | **39,134** |

n and N interviewed as proxies

| | Pre-Pandemic | Pandemic | Total |
|---|---|---|---|
| n | -1,788 | -1,109 | **-2,897** |
| N | | | **-1,111** |

Eligible n and N over 18y old:

| | Pre-Pandemic | Pandemic | Total |
|---|---|---|---|
| n | 65,785 | 84,455 | **150,240** |
| N | | | **38,146** |

n and N over 18y old:

| | Pre-Pandemic | Pandemic | Total |
|---|---|---|---|
| n | -1,864 | -905 | **-2,769** |
| N | | | **-988** |

Eligible n and N with non-missing cross-sectional weight

| | Pre-Pandemic | Pandemic | Total |
|---|---|---|---|
| n | 36,005 | 69,246 | **105,251** |
| N | | | **26,053** |

n and N with missing or zero weight

| | Pre-Pandemic | Pandemic | Total |
|---|---|---|---|
| n | -29,780 | -15,209 | **-44,989** |
| N | | | **-12,093** |

Eligible n and N with non-missing values in the outcome variable:

| | Pre-Pandemic | Pandemic | Total |
|---|---|---|---|
| n | 35,607 | 66,704 | **102,311** |
| N | | | **25,684** |

n and N with missing values in the outcome variable:

| | Pre-Pandemic | Pandemic | Total |
|---|---|---|---|
| n | -398 | -2 542 | **-2,940** |
| N | | | **-369** |

Eligible n and N in employment:

| | Pre-Pandemic | Pandemic | Total |
|---|---|---|---|
| n | 24,311 | 38,447 | **62,758** |
| N | | | **15,476** |

n and N not in employment:

| | Pre-Pandemic | Pandemic | Total |
|---|---|---|---|
| n | -11,296 | -28,257 | **-39,553** |
| N | | | **-10,208** |

Eligible n and N with no missing values in key variables*:

| | Pre-Pandemic | Pandemic | Total |
|---|---|---|---|
| n | 21,841 | 33,505 | **55,346** |
| N | | | **13,721** |

*: Outcome, exposures and all covariates used in Models 1 and 2

n and N with missing values in our key variables:

| | Pre-Pandemic | Pandemic | Total |
|---|---|---|---|
| n | -2,470 | -4,942 | **-7,412** |
| N | | | **-1,755** |

Eligible n and N who participated in wave 9 and at least one Covid-19 survey:

| | Pre-Pandemic | Pandemic | Total |
|---|---|---|---|
| n | 12,192 | 29,015 | **41,207** |
| N | | | **6,474** |

n and N that only participated in either pandemic or Covid-19 surveys:

| | Pre-Pandemic | Pandemic | Total |
|---|---|---|---|
| n | -9,649 | -4,490 | **-14,139** |
| N | | | **-7,247** |

Eligible n and N with no missing values in the occupation variable:

| | Pre-Pandemic | Pandemic | Total |
|---|---|---|---|
| n | 8,210 | 18,900 | **27,110** |
| N | | | **4,485** |

n and N with missing values in the occupation variable:

| | Pre-Pandemic | Pandemic | Total |
|---|---|---|---|
| n | -3,982 | -10 115 | **-14,097** |
| N | | | **-1,989** |

Figure S1. Stratified Regression models (UK Country of residence)

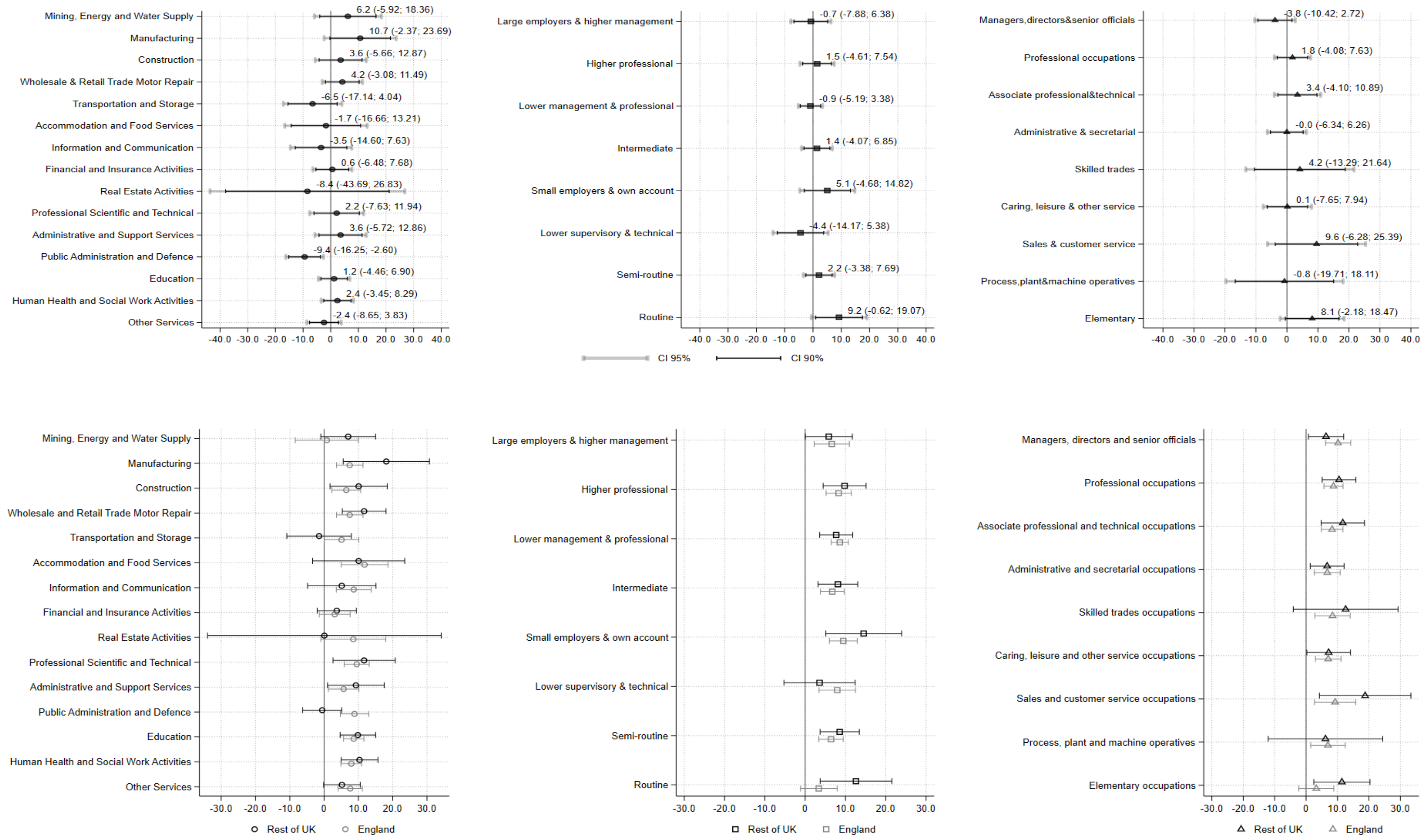


Note: circles represent industries, squares social class and triangles occupational groups

Figure S2. Stratified Regression models (Age)

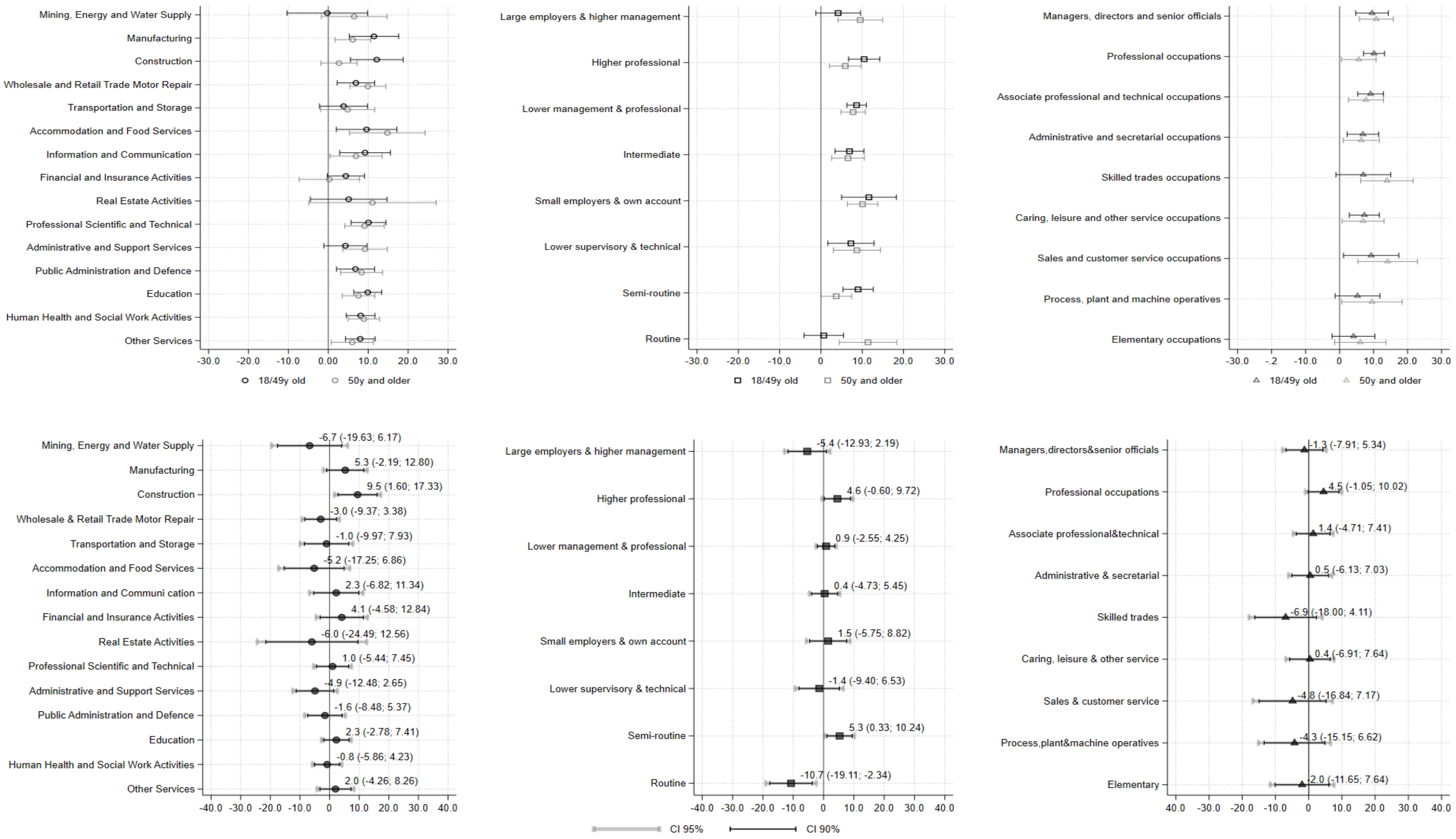


Note: circles represent industries, squares social class and triangles occupational groups
